# A Scoping Review of Studies on Secure Messaging through Patient Portals: Persistent Challenges and Potential Solutions

**DOI:** 10.1101/2025.05.19.25327942

**Authors:** Yawen Guo, Di Hu, Yiliang Zhou, Tianchu Lyu, Ziqi Yang, Tera L. Reynolds, Kai Zheng

## Abstract

Secure messaging (SM) has seen increasing adoption over the past decade, prompting growing interest in understanding its impact on healthcare delivery. This review examines key themes in the existing SM research, such as usage patterns, perceived benefits, and persistent challenges, to identify research gaps and inform opportunities for sociotechnical solutions in the artificial intelligence (AI) era that could enhance patient–provider communication effectiveness. Searches were conducted in PubMed, Scopus, IEEE Xplore, ACM Digital Library, Cochrane CENTRAL, CINAHL, and Web of Science. Following the PRISMA guideline, we identified 366 relevant peer-reviewed studies published from January 2009 to September 2025. Existing research has primarily investigated (1) the effect of SM use on clinical outcomes, (2) adoption patterns, and (3) user experiences. The literature shows that SM promotes patient engagement, care coordination, and patient–provider communication. However, significant challenges persist, including privacy concerns, limited access for vulnerable populations, patient misuse, and increased clinician burden. Educational initiatives and patient-centered design are essential for promoting appropriate and accessible use of SM. Emerging AI solutions also show promise in enhancing SM use, particularly for message triaging and drafting replies. The integration of such AI solutions must be guided by robust governance frameworks to ensure transparency, maintain trust, and align with evolving clinical, billing, and regulatory environments. Future research should include more diverse care settings and populations, prioritizing the development of equitable sociotechnical tools and interventions that can be seamlessly integrated into clinical workflows.

## Introduction

The Health Information Technology for Economic and Clinical Health (HITECH) Act has promoted the meaningful use of electronic health records (EHRs) to enhance healthcare quality, patient engagement, and data security[1]. Under its Meaningful Use provisions, secure electronic messaging was established as the standard for patient-provider communication and was promoted in support of EHR innovations like telehealth to enhance care between visits [2–4]. Secure messaging (SM), defined as “any electronic communication between a provider and patient that ensures only those parties can access the communication”, is a core feature of patient portals and plays a critical role in the broader landscape of telehealth [5–8].

SM has become a widely adopted feature in patient portals, supporting patients in self-managing health conditions and coordinating care[9,10], showing potential to improve patient engagement and provider workflows via enabling multimodal asynchronous information exchange[11]. Despite its growing use, challenges persist in expanding adoption, ensuring effectiveness, and addressing overuse, particularly since the COVID-19 pandemic [12–18]. As SM has taken on greater importance in clinical care, policy responses have followed. For example, CMS introduced billing codes for portal messages that involve medical decision-making and require more than five minutes of clinician time within a seven-day period[11]. These policies acknowledge the clinical value and resource burden of SM while underscoring the need for ongoing evaluation of their impact on care delivery, equity, and provider workload [15,19].

Machine learning (ML) and deep learning(DL) approaches have long been explored to enhance SM in support of a variety of clinical tasks, including triaging, message classification, and workload prediction[20–22]. More recently, advances in generative AI (GenAI) and large language models (LLMs) have prompted EHR integration of these technologies for message drafting and workflow optimization[23–25], particularly with respect to reduction of provider workload associated with SM[26–29]. Despite considerable enthusiasm, the real-world benefits of such AIML-enhanced SM approaches have yet to be firmly established. Such an evaluation presents a two-fold challenge: first, elucidating how SM affects care delivery and clinical workflows, and second, determining the additional impact of AIML on these processes.

While several SM studies exist that provide important insights on the former point, many were conducted before the widespread adoption of modern EHRs and digital health technologies and thus reflect an outdated technological landscape. Early reviews examined its potential to enhance patient-provider communication, focusing on user expectations, confidentiality concerns, and technological limitations reflective of the pre-2010 landscape[30–32]. Conversely, more recent reviews highlight specialized aspects rather than a generalized view of the field, covering specific vendor applications [33], disease- or population-specific implementations[34–36], or possessing computational or methodological foci[37,38]. A comprehensive understanding of SM’s role in care delivery—its benefits, limitations, and areas of adoption—is essential for determining whether, where, and how AI should be integrated into clinical workflows.

In this article, we therefore aim to systemically understand the current and prospective role of the patient–provider SM communication in clinical care: its benefits, drawbacks, and implications for potential solutions. In doing so, we establish a contemporary foundation to identify opportunities for, and risks inherent to, adoption of AIML-enhanced SM from the perspective of the health system as a whole. We then conclude with further elaboration on several such identified opportunities and risks.

## Results

A search across seven databases yielded 1327 unique studies. A total of 773 studies were removed as they were unrelated to SM after title and abstract screening. After full-text screening of 554 articles, 188 were excluded based on predefined criteria (Figure 1). We included 366 relevant studies in this review. The process followed PRISMA guidelines to ensure methodological rigor and comprehensive coverage. Agreement between reviewers was substantial at both the title and abstract screening phase (κ = 0.67) and the full-text review phase (κ = 0.66). All conflicts were resolved through consensus.

**Fig. 1.**
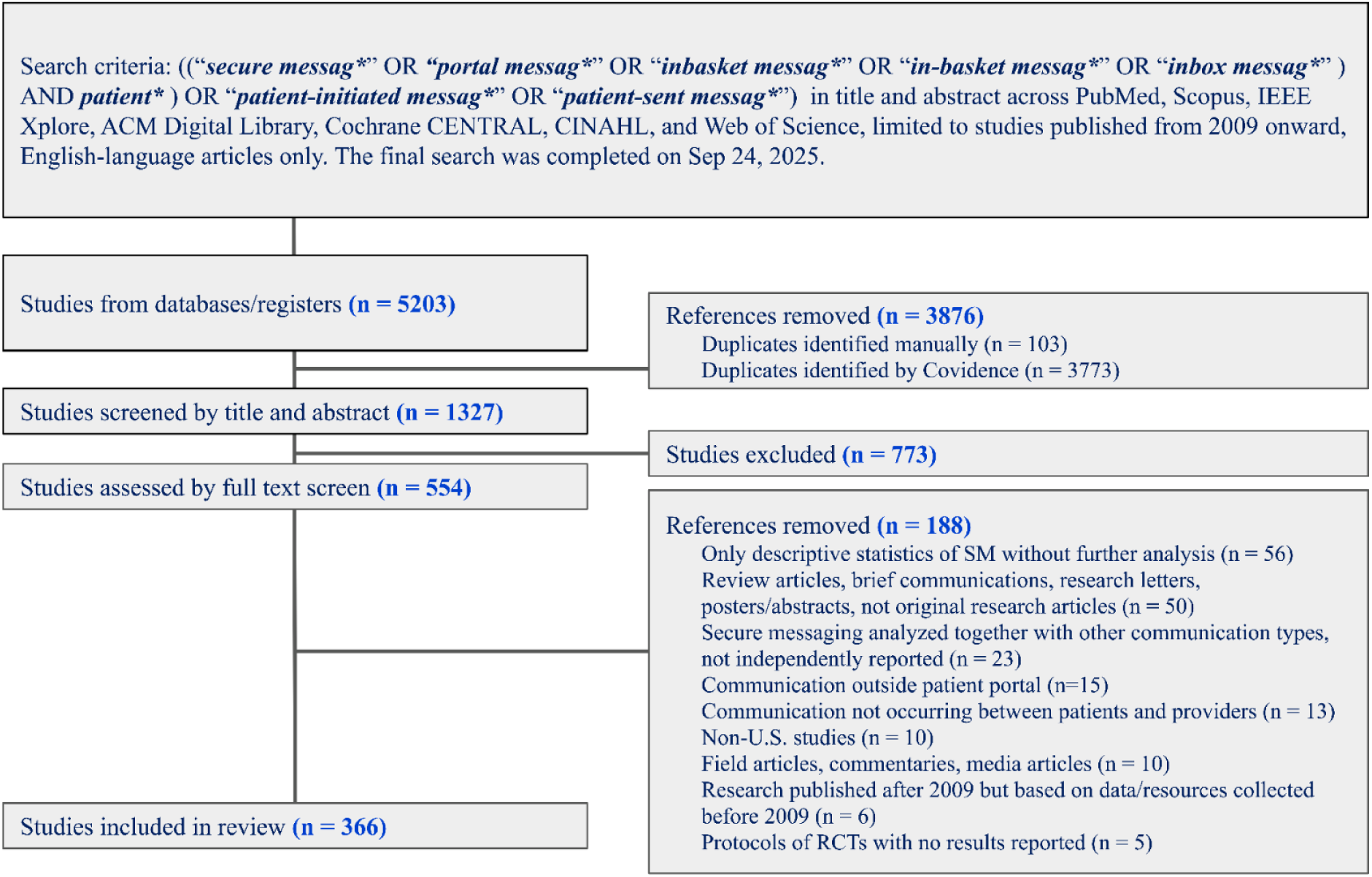
Literature search and selection process.

Our analysis categorized these 366 studies into nine distinct research themes (Figure 2), capturing a wide range of topics related to SM within EHRs. The following sections present findings from each category, highlighting key insights on healthcare communication, patient engagement, clinical outcomes, and technological developments.

**Fig. 2.**
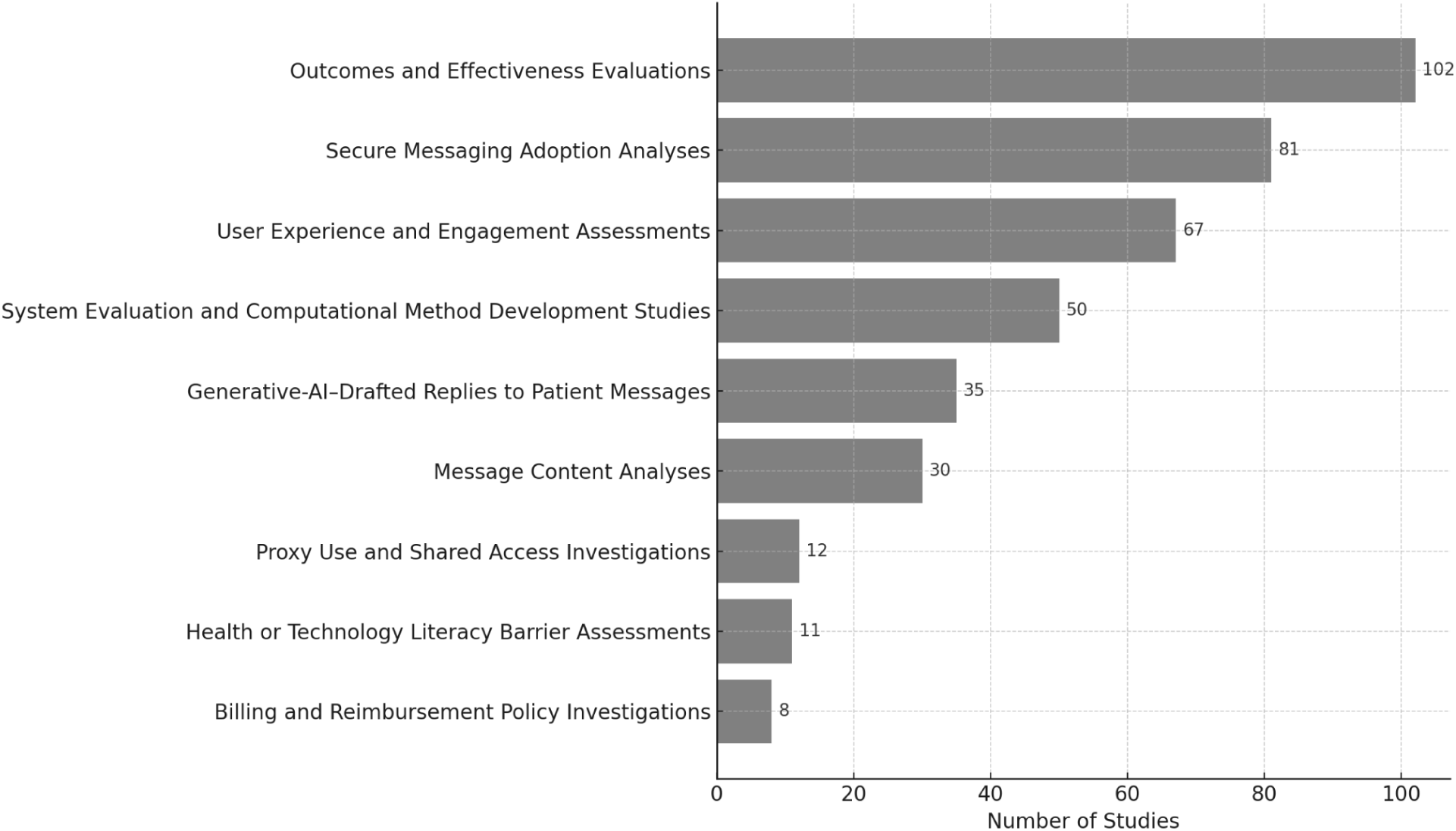
Distribution of Research Themes Across Reviewed Articles (n = 366) *\* Note: Some studies were coded under multiple categories*

### Outcomes and Effectiveness Evaluations (n=102, 27.9%)

Most studies accessing outcomes or effectiveness showed that SM utilization is associated with benefits for patient care and patient, clinician, and researcher work across various clinical conditions, care settings, and patient populations. For example, SM has been found to enhance diabetes care and self-management[39–48], and improved survival outcomes and symptom monitoring among oncology patients[49,50]. Use of SM has been associated with improved hypertension management via better blood pressure monitoring and medication intensification[51–53].

Studies assessing the effectiveness of SM outreach and interventions have been overall positive, but somewhat mixed. 12 studies have linked SM with increased rates of vaccination[54–59], colorectal and hepatitis C screenings[60–62], pediatric care attendance[58,63,64], and genetic testing for Alpha-1 Antitrypsin Deficiency[65]. Interventions conducted via SM also showed potential association with improved medication adherence among patients, including for smoking cessation[61–65], chronic myeloid leukemia[66], spina bifida[67], and depression[68]. In surgical contexts, simplified dismissal instructions after prostatectomy were reported to be associated with fewer postoperative patient-initiated messages and ED visits[69]. Among elderly patients, SM communication has been found to facilitate care planning and advance directives completion[70]. In pediatric young adult settings, SM has been reported to support medical decision-making[71], deliver educational interventions in suicide prevention and HIV care[72–76], and facilitate follow-ups following office-based circumcision[77,78].

Secure messaging related usage metrics has been used as an indicator of hospitalist efficiency and patient portal engagement[79–83]. It has been reported to influence clinical workflow by increasing patient engagement and supporting the adoption of e-visits[84–91].However, a higher volume of SM interactions was reported to reduce new patient intake capacity[84,92]. While its association with emergency visits varies by conditions[93–95], SM has generally been linked with more efficient primary care workflows, including lower urgent care utilization and fewer unnecessary visits[96–100]. Studies have found that provider burnout remains a concern because of large message volume[101], while message characteristics, such as sentiment scores, do not appear to be a major contributing factor[102]. SM has also demonstrated effectiveness as a clinical research recruitment tool, achieving high response rates compared to mail, phone, and social media methods across diverse clinical trials[103–112].

Studies comparing SM with traditional communication methods highlight varied user preferences influenced by clinical context and demographics, suggesting that tailored communication approaches are essential[109,113–124]. Studies report lower response rates of SM for COVID-19 research recruitment and preventive screening when compared to traditional in-person, phone, or mail communication[113,114,125,126]. Influenza vaccination studies show mixed results, with some reporting associations with higher uptake[117,127,128], while others find no population-level benefits from SM reminders[129,130] or content-tailored SM[131]. Although certain studies have suggested benefits of SM interventions, others report no significant advantage of SM outreach for smoking cessation[74,132,133] or HIV medication refills[134,135]. Similarly, a randomized trial found that a single SM was not associated with an increase in HbA1c follow-up testing[136].

### Secure Messaging Adoption Analyses (n=81, 22.1%)

Secure messaging adoption has been expanding substantially since its introduction around 2010 and is now reported as the most used patient portal function[137–140]. During this time, most office-based physicians adopted SM, although uptake was lower among solo and non-primary care providers[141–143]. Disparities emerged following federal incentives, with certified health IT use positively associated with SM adoption, as well as providers’ expectations that SM would improve performance and their overall attitudes toward its use[144–146]. Research shows SM volume continues to rise but per capita rates have plateaued, indicating SM adoption is still expanding [143,147–153]. The COVID-19 pandemic triggered a major surge in use, and persistent high volume has been suggested to contribute to multitasking, workflow disruptions, and potential medical errors[154–163]. A national study found that only 1.9% of veterans used SM during early adoption, with uptake varying by condition and demographics[164].

Patient SM usage patterns vary across demographics. Patient users are more often younger [89,165–173], women [166,169,171,174–178], Caucasian [89,166,168,169,175,177,179,180], married [89,107,168,169,177,181], employed [89], privately insured [89,145,171,182,183], and residing in high socioeconomic neighborhoods [89,144,167,184] with higher education levels[165,169,185,186]. Many also have poor physical[165] or mental health conditions[175,187,188], and higher levels of morbidity or comorbidity[167–169,171,188,189]. Active patient users are generally English speaking[177,180,190], with high EHR usage and more outpatient visits[191–194]. Portal refresh behaviors also shape adoption patterns, with patients who more frequently refreshed test results more often initiating SM[195]. Among adolescent patients, SM users were more often female or transgender with prior portal experience[196]. In contrast, lower SM engagement has been observed among underrepresented minorities[140,145,197–199], veterans experiencing homelessness[187,200,201], individuals with limited English proficiency or low health literacy[189,202–205], and patients facing poor internet access and other digital disparities[167,175,185,206,207].

Patient–provider interactions shape SM adoption, and have been described as being influenced by behavioral health models, clinical roles, and the characteristics of both patients and providers[208,209]. Female patients have been reported to be more likely to request medical guidance via SM but receive fewer confirmations in response[210]. Female physicians are more often than male physicians to send and receive SM, which may in turn contribute to higher rates of burnout among female providers[174,211–214]. However, among pediatric surgical specialists, a recent analysis found no significant differences in inbox message volume or message length after adjusting for case complexity, suggesting that gender disparities in messaging may be context-dependent and vary by specialty[215].

### User Experience and Engagement Assessments (n=67, 18.3%)

Studies evaluating user experiences report high satisfaction among patients [216–220], and strong enthusiasm from providers[221,222], particularly in relation to medication refills, appointment scheduling, test result access, clinical inquiries[14,223–230]. Patients view SM as a tool that complements face-to-face communication, especially regarding sensitive issues like sexually transmitted diseases(STDs) and erectile dysfunction[216,231,232]. SM use has also been reported to improve perceived access to care[14,223,231,233–236] and to strengthen patient-provider relationships[237]. Providers have described valuing SM for its potential role in improving workflow, care coordination, and enabling remote work[233,238].

However, patients have raised concerns about provider burden from uncompensated SM interactions, delayed responses, unclear messages, and difficulty reaching specific providers–issues that may contribute to tracking challenges and inconsistent communication[12,226,239–245]. Providers have likewise reported increased workload due to time-intensive message management[235,243,244,246–253], and stress from inappropriate or emotionally challenging messages[12–14,226,254]. Findings from several studies suggest that message-related work created additional burdens, such as constant redirection of attention, lengthy inbox management, and heavier mental workload—factors that have been associated with reduced efficiency and responsiveness[246,249,255–258]. A recent study of over 1,700 physicians reported that patient messages requesting medical advice were associated with significantly increased after-hours work, with specialists experiencing a greater burden than primary care physicians[259]. Additional concerns noted in the literature include liability risks[248], limited effectiveness of SM in inpatient settings[260], and inadequate training among medical trainees in using SM[261].

User-perceived barriers to SM adoption include patient misconceptions and previous negative experiences—such as uncertainty about what counts as a ‘non-urgent’ message or concerns about whether physicians have sufficient time to devote to messaging[141,231,262]. Patients have also reported challenges related to message ambiguity and complexity, particularly among elderly patients with lower literacy[141,238,263]. Other barriers described in the literature from the provider perspective include delegation of messaging tasks that complicate communication[13], limited technology and internet access[13,206,225], and unclear guidelines for messaging use[141,264,265]. Further work has similarly characterized inbox tasks as fragmented and labor-intensive, often requiring additional articulation work to clarify conflicting or redundant requests[266]. One study reported that the use of medical scribes was not associated with reductions in time to completion of SM[267]. Proposed solutions emphasize education and systematic training to instill more efficient inbox management habits of providers [234,241,255,268–272], accessible user interfaces with clear messaging instructions[141,257,264], and AI-driven tools designed to improve patient literacy and support physician workflows[141,273,274]. Additional recommendations include strategic message management practices such as team-based message sharing[226,234,243,255,275], crafting clear and empathetic messages[276,277], and standardizing administrative processes to enhance efficiency and reduce clinician burden[269].

### System Evaluation and Computational Method Development Studies (n=50, 13.7%)

Early evaluations of SM systems reported rapid consultations turnaround times during pilot e-visit deployments[278]. However, later studies identified persistent system-level usability and access challenges including delayed message response times, suggesting limitations in platform responsiveness and interface design[279,280]. System analyses using signal and log data highlighted rising message volumes and uneven clinician workload distribution, with primary care physicians and nurse practitioners handling most communication during standard work hours[170,281]. Usability research in hospital settings identified functionality gaps, such as clinical document sharing errors[282], while broader evaluations linked increased SM use to patient access to EHR data[283].

Recent studies leverage patient–provider SM data to improve clinical efficiency and communication using computational methods. Theory-informed tools—designed to enhance digital engagement, support content creation, and implement feedback mechanisms—were associated with increased portal use among users with diverse literacy and numeracy skills[277,284–286]. Automated assessments of readability and health literacy have provided insights into message complexity, informing both clinical care and population-level interventions[287–291]. Framework-based approaches include SPICE, which emphasizes support, partnership, and information-giving, and a Chain-of-Thought routing framework leveraging LLMs; both illustrate structured strategies intended to improve message handling and patient-centered communication[234,292]. An extended Technology Acceptance Model that included affordances and communication efficacy found that young adults’ intentions to use portal messaging were positively associated with editability and communication efficacy, but negatively associated with persistence[293].

Rule-based, natural language processing(NLP), deep learning(DL) models have been developed to triage SM by risk and urgency[22], classify information type (e.g., clinical, medical, logistical, or social)[20,21], detect medical decision complexity[26–29], and identify caregivers and caregiving network among people living with dementia[294]. A prospective deployment of a fine-tuned NLP classifier for automated routing reported reduction in response times and staff interactions while maintaining high accuracy across message categories[295]. They were also applied to identify primary patient concerns, social needs, and mental health crises[296–306]. Efforts to extract actionable insights from messages include building FHIR(Fast Healthcare Interoperability Resources)-aligned data models to standardize SM content and enable downstream NLP analyses, integrated semantic and contextual features for improved categorization, and combined message-based features with predictive models to support clinical decision-making[307–310]. Additionally, vector-based representation techniques have been explored with a focus on optimizing word embeddings for small, domain-specific datasets[311], and prompt engineering has been used to generate synthetic drug-related messages to reveal language patterns[312]. More recently, studies have applied LLMs to tasks such as triaging patient messages to appropriate clinics or urgency levels, detecting knowledge questions, tailoring patient support tools for chronic disease management, and identifying emergencies in real time[313–317].

### Generative-AI–Drafted Replies to Patient Messages (n=35, 9.6%)

A rapidly growing body of 35 studies published between 2023 and 2025 has examined the use of large language models (LLMs) to generate draft responses to patient portal messages. Live implementations in EHR pilots occurred mainly in Epic in-basket workflows, often pairing GPT-4 with institutional customization[23,301,318–322], while the majority of studies used simulated or de-identified message sets across diverse specialties[24,25,323–339].

AI-generated replies were often rated as comparable to, or in some cases better than, clinician-authored responses in terms of informational adequacy and clarity, and several studies highlighted enhanced empathy, particularly in GPT-4 outputs[25,319,324,330–333,340–342]. Model benchmarking further reported GPT-4’s consistent outperformance of earlier LLMs across domains[323,328]. Evidence from real-world pilots showed low adoption of draft functions and near-universal need for clinician edits before sending. Drafted messages were generally longer than clinician-authored replies, and objective time savings were inconsistent—though many clinicians perceived reduced burden[23,318–320,343]. Metrics such as edit distance and message length were commonly used to quantify these patterns[339]. Several studies emphasized that iterative prompt engineering and contextual grounding—for example, incorporating the most recent assessment and plan—were associated with improvements in draft quality and clinician acceptance[318,328,334,343,344]. Human-in-the-loop prompt refinement and structured prompt templates were also reported as effective strategies[321,335].

Clinicians described drafts as useful starting points that could reduce cognitive load, though some expressed concerns about over-reliance[257,325,330,332]. Patients in blinded evaluations frequently favored AI replies for empathy and readability[319,327,330], while mixed feedback emerged around the tone of lengthy or formal responses[324,326]. Risks were documented across multiple contexts, including hallucinations, incoherence, and misinterpretation of clinical nuance[25,323,326,333,336,345]. These findings underscore the necessity of clinician oversight in all implementations[331]. Participants across several studies were often unable to reliably distinguish AI-authored from human-authored replies[327,330]. While disclosure of AI involvement slightly reduced patient satisfaction, transparency was widely viewed as necessary for trust and accountability[325,334,346,347].

### Message Content Analyses (n=30, 8.2%)

Studies analyzing SM content have used both qualitative and computational methods to examine communication patterns. Common topics include medication-related issues, appointment scheduling, lab results, referrals, and other administrative tasks[39,39,115,156,166,348–357]. Patients have been observed to use SM to report symptoms, express psychosocial concerns, seek medical advice, and convey appreciation or complaints[115,156,166,176,351,352,354,358–363]. SM content also reveals communication around sensitive health topics such as STDs and cannabis use[216,364]. In surgical populations, SM content has focused on drains, wounds, pain, and activity restrictions, with message patterns varying by patient age and mental health status[365]. Tone analysis reveals that most messages are neutral and respectful, though cyber-incivility and problematic language occasionally challenge professional standards[254,366,367]. Evidence suggests that provider replies may include less language that fosters partnership or emotional support[368], and provider-initiated messages often reflect greater clinical complexity[369]. Recent work has also applied large-scale NLP to patient portal messages to surface patient-driven research priorities in cancer care, with approximately one-third of the AI-generated topics rated by clinicians as highly meaningful and novel[370].

### Proxy Use and Shared Access Investigations (n=12, 3.3%)

This body of research focuses on patient portal use among adolescents with their guardians, and elderly patients with their caregivers. Studies have highlighted benefits such as convenient communication and patient empowerment, but also raised concerns about privacy and confidentiality—particularly for adolescents under the Cures Act Final Rule[371,372]. Analyses indicate that more than half of messages from adolescent accounts are accessed by guardians, underscoring privacy management challenges[373,374]. For elderly populations, research has consistently reported caregiver use of portals and SM as proxies, reflecting significant reliance on caregiver support[357,375–377]. Caregiver proxies have been observed to more commonly manage portals for older adults, males, non-white patients, non-English speakers, noncitizens, and those with public insurance[378]. To address these challenges, recent studies developed computational methods to identify non-patient authors, aimed at improving management efficiency and strengthening privacy safeguards in patient portal communications[189,379,380].

### Health or Technology Literacy Barrier Assessments (n=11, 3.0%)

Several studies have emphasized the importance of health and digital literacy for the adoption and effective use of SM. Research indicates that while patients often write in simpler language, a mismatch in communication complexity has been observed in a notable portion of exchanges[287]. Studies have also identified disparities linked to the digital divide, highlighting barriers such as low digital literacy[140,203]. Providers have reported technology-related difficulties, including complex login procedures and frequent log-offs[12]. Investigations into SM complexity suggest that automated feedback mechanisms may support message clarity and user education, thereby helping providers communicate more effectively[277]. Computational models using linguistic analysis of patient messages have also shown promise in assessing health literacy and identifying educational needs automatically[238,277,288–291,381].

### Billing and Reimbursement Policy Investigations (n=8, 2.2%)

Recent studies examining billing policies for SM e-visits have highlighted changes in clinician billing behaviors and message usage patterns following the 2020 decision by CMS to allow billing for SM. Initial assessments reported modest declines in message volume after CMS’s decision and low physician participation in billing activities[15,19,382–384], even though providers generally accepted the new model[384]. Emerging research has proposed refining billing strategies using predictive models based on clinical complexity rather than time spent, with the aim of improving billing accuracy and effectiveness[29,385]. Investigations in specialty areas, such as ophthalmology, have reported a temporary increase in billing during the pandemic, underscoring challenges in sustaining equitable SM billing across diverse healthcare contexts[386].

## Discussion

This scoping review systematically maps the evolving landscape of SM research, emphasizing its role in patient–provider communication and the increasing influence of AI in shaping its implementation. SM has shown clear benefits in enhancing access, continuity, and care coordination. However, challenges around equity, usability, and provider burden persist, underscoring the need for coordinated efforts among all key stakeholders, including patients, providers, researchers, and policymakers. To guide future work, we discuss findings across four major domains, reflecting the research themes identified in our results.

### Secure Messaging Supports Care but Evidence Across Populations and Care Settings Remains Limited

Across the included studies, the majority has been conducted within single healthcare institutions, particularly emphasizing primary care settings. Investigations often target specific populations: e.g., older adults and veterans[101,245,357,375], or patients with specific diseases, such as chronic illnesses[240,354,380]. Concurrently, there has been limited exploration of SM’s benefits and limitations beyond primary care, such as in emergency or specialty settings. While the bulk of SM’s applicability is in primary care[280,287,303,346], they may also be useful in specialty settings[95,309]. These investigations on SM systems in limited, focused, settings inherently also limit the broader applicability and generalizability of findings. Future research should therefore diversify study populations, broaden the range of health conditions examined, and include varied care settings. Future research should therefore diversify study populations, broaden the range of health conditions examined, and include varied care settings.

### Patient Misconceptions, Equity Barriers, and Privacy Risks Limit Adoption

Although SM offers important opportunities to strengthen patient–provider communication[30,31,34], many patient-side barriers persist, including misconceptions about appropriate use, inequities in adoption, and privacy concerns.

#### Patient Misconceptions, Privacy Concerns, and Misuse of SM

Patients often hold misconceptions about SM, influenced by negative experiences with telehealth, uncertainty about who receives their messages, and concerns over privacy and system responsiveness[141,231]. Some patients hesitate to use SM due to fear of overburdening their providers, while others misuse it for urgent needs[240,242]. Although computational methods such as urgency classification have been developed to support triage[351], they alone are insufficient to ensure appropriate use. AI-based tools have the potential to guide patients in real time by suggesting appropriate message categories, clarifying urgency, or simplifying language. However, these solutions must be paired with user education to be effective, as patients need guidance on when and how to use SM appropriately[12,226,241,244]. Some health systems have begun embedding brief guidance within patient portals or visit summaries, yet few efforts are tailored to individuals with limited digital literacy, language barriers, or disabilities. Furthermore, little is known about the effectiveness of these approaches or whether they reach the patients most in need. Clear communication at the point of care, e.g., setting expectations about appropriate message use during visits, may help address these misunderstandings without adding new technological burden. Finally, additional safeguards must be in place to protect patient privacy when messages are accessed or sent by non-patient senders, such as caregivers, guardians of adolescents, or individuals assisting patients with disabilities[357,372,373,387]. Potential strategies include role-based access controls, clear proxy account policies, and automated alerts to help patients maintain awareness of who can view or send messages on their behalf.

#### Expanding Access and Equity Through Patient-Centered Design

Non-English speakers and the elderly continue to face technical barriers that limit their effective use of SM[189,378]. Addressing these challenges requires patient-centered design that incorporates user-friendly interfaces and supports low-literacy communication to enhance functionality and accessibility for diverse populations[366,368,388–391]. Individuals from low socioeconomic backgrounds often face barriers to accessing SM, particularly when care is delivered across multiple providers[89,144,167]. Current interventions are often passive, relying on patient-initiated engagement, which may inadvertently exclude underrepresented groups who are unaware of or hesitant to use SM. While some outreach efforts exist, such as reminders for vaccinations or preventive screenings, few are tailored to the needs of specific populations. Future directions include co-designed educational materials, multilingual and plain-language prompts, and accessibility features, alongside rigorous evaluation of their impact. AI-powered tools may further support equity by delivering simplified language, visual aids, and personalized educational content to improve comprehension and engagement.

### Provider Burden Reflects System and Policy Gaps

Beyond patient-facing barriers, our review identified substantial strain on providers, driven by increasing message volumes, unclear workflows, and limited reimbursement mechanisms.

#### Reducing the Burden on Providers

Misuse of SM for urgent issues can shift liability and clinical responsibility to providers, potentially compromising patient care[254,367]. Patients’ use of inappropriate language or emotionally charged messages may contribute to a stressful professional environment and emotional exhaustion[254,367]. Beyond emotional strain, operational burdens include frequent attention shifts, prolonged inbox management, and increased cognitive load, which can reduce the quality and timeliness of responses[246,249,255]. The surge in message volume further intensified these pressures, often extending work into personal time and increasing physician burnout[226,243,258,392,393]. Disparities in SM use between physician- and hospital-owned practices suggest that organizational structures also shape provider engagement[179]. To address these challenges, some interventions have introduced structured workflows or support systems, such as team-based inbox management or message triage protocols, but few studies have systematically evaluated their effectiveness in reducing workload or improving care delivery. There is also a role for provider-facing education to ensure consistent messaging around SM use during clinical encounters[234,241,255,268–272]. Future research should assess the comparative effectiveness of these workflow interventions and identify conditions under which AI-driven support, such as automated triage or message summarization, can reduce workload without undermining clinical judgment. Organizational strategies such as clearer role delineation and protected time for messaging are critical components of reducing provider burden. However, AI-generated drafts may introduce new forms of cognitive and time burden, underscoring the importance of human-centered workflow design to ensure net workload reduction[394]. A balanced approach that integrates automation with clinician-centered workflow design is needed to alleviate provider burden.

#### Billing Models and Economic Considerations in SM

While early billing policies for SM focused on clinician time and decision-making complexity, ongoing implementation has raised broader concerns around equity, resource allocation, and communication dynamics[15,19]. Modest physician engagement and reduced patient messaging suggest hesitancy toward new billing protocols and potential shifts in patient behavior[382,383]. Predictive modeling based on clinical complexity has offered strategies to align care delivery with billing practices[385]. Although AI-enhanced billing tools could theoretically analyze message content and estimate clinical effort, they remain largely untested[395]. As these technologies evolve, it is important to monitor their impact on provider documentation and patient access. Future research should further evaluate how providers and patients adapt to SM billing and explore how AI can contribute to transparent and sustainable reimbursement models.

### AI-Augmented Solutions Offer Promise but Require Careful Governance

Computational and AI-driven approaches are increasingly being explored to manage message volume and workflow, improve literacy support, and generate draft replies.

#### Computational Method for System Support

For current computationally augmented SM implementations, the primary focus has been on automated triage and response prioritization. Studies have applied approaches ranging from traditional ML models such as logistic regression, support vector machines, and random forests[21,22,26] to DL architectures, particularly BERT and its variants[300,396]. These methods have been used to classify message urgency and decision complexity[302,379,397], and to assess readability or literacy mismatches in patient–provider communication[288–290]. While most work has been limited to retrospective analyses or small-scale pilots, findings suggest promising utility for identifying high-risk messages, enhancing literacy support, and improving workflow efficiency. Future directions include adapting these tools for live clinical workflows, integrating them with structured EHR data, and examining their impact on equity (e.g., whether automated triage inadvertently deprioritizes messages from underserved populations). As LLM-based methods are now being applied to similar SM tasks[313–317], future research should prioritize rigorous evaluation of their accuracy and safety, and determine how they can complement existing ML and DL approaches[37].

#### Integrating Generative AI in SM Workflows Requires Careful Research and Governance

The integration of GenAI and LLMs into SM offers new opportunities to improve efficiency and reduce clinician burden, especially in the management of in-basket messages[390,398]. While providers often prefer human-generated messages for their readability and empathy[25,319,339], patients are generally receptive to GenAI-generated messages, especially when tone and context are appropriately managed[329]. Recent research has begun exploring GenAI’s feasibility across varied message types and clinical scenarios to improve applicability and trust in message drafting[325,331,344,399]. Pilot studies are also testing GenAI for triage, response generation, and workload redistribution within SM systems[23,318]. While early studies have explored GenAI feasibility, widespread integration will require more robust evaluation of message quality and broader impacts on communication safety, patient–provider relationships, and workflow efficiency. As SM continues to evolve, human-centered governance will be essential to ensure that GenAI enhances care while maintaining trust and accountability.

### Limitations and Future Work

This review included only full-length archival articles on empirical studies, excluding brief reports, research letters, and industry case studies that may have provided early insights into emerging trends. Future reviews should consider these formats to broaden analytical scope. Our search approach focused on SM terminology common in portal-based systems, which may have excluded earlier studies using different descriptors such as “electronic messages”. We also limited inclusion to patient–provider communication via portals, excluding studies on stand-alone SM applications and interprofessional communication (e.g., physician–nurse exchanges) to maintain a clear focus. Future research should examine SM in alternative platforms and team-based care contexts to better capture its broader impact on healthcare communication and coordination.

## Conclusion

This scoping review synthesized literature on patient–provider secure messaging (SM) within patient portal, highlighting key themes including user perspectives, communication patterns, clinical outcomes, and technology adoption. While SM offers clear benefits for patient engagement and care coordination, challenges persist around provider workload, digital access, and equitable implementation. AI-assisted tools show promise in addressing these barriers but must be paired with thorough evaluation and careful integration into clinical workflows. We underscore the need for user-centered system design, targeted education to improve digital and health literacy, and informed policy development regarding billing and physician burden. Future research should further explore how LLMs and GenAI can be responsibly applied to support secure and efficient patient–provider communication.

## Materials and Methods

### Literature Search and Selection

We conducted a literature search across PubMed, Scopus, IEEE Xplore, ACM Digital Library, Cochrane CENTRAL, CINAHL, and Web of Science. The final search strategy combined the terms (“secure messag*” OR “portal messag*” OR “inbasket messag*” OR “in-basket messag*” OR “inbox messag*” OR “patient-initiated messag*” OR “patient-sent messag*”) AND patient* in the title and abstract, restricted to studies written in English and published from 2009 onwards. A detailed breakdown of search strategies for each database, including syntax variations and filters, is provided in the Supplemental Materials Table S1. The search process was conducted in four waves to ensure comprehensive coverage of the literature over the study period. The first search was completed in December 2023 and provided the initial corpus used to pilot and refine the inclusion and exclusion criteria. A second and third search were conducted in October 2024 and April 2025 to update the corpus and incorporate new publications that had appeared in the interim, while also allowing refinement of coding practices and terminology. The final search was completed in September 2025, ensuring that the review reflects the most recent body of evidence. Results in this manuscript are based on the combined set of studies identified across all four searches. The titles and abstracts of the identified articles were independently screened by YG and another author (DH, YZ, TL, or ZY). Articles identified as relevant after the title and abstract screening process then underwent a full-text review to determine final inclusion(YG and DH). Any disagreements were resolved through consensus meetings with a third author (KZ). Inter-rater reliability was assessed using Cohen’s κ in Covidence for both the title and abstract screening stage and the full-text review stage. Because all records from different search waves were uploaded together, the κ values represent overall agreement across the full screening process.

We included original, peer-reviewed full-length empirical studies in which patient–provider SM was a central focus. We excluded studies that: (i) did not examine SM directly, (ii) analyzed communication solely between providers, (iii) combined SM with other modalities (e.g., telephone or email) without separate reporting, (iv) were conducted outside the U.S., or (v) reported only basic descriptive metrics of SM use (e.g., counts or enrollment) without analytical depth. For example, studies reporting only annual message volumes without linking them to outcomes, content, or user experience were excluded.

### Data Extraction

To formulate the codebook, YG and DH iteratively coded sets of 10 articles, followed by consensus discussions with KZ to refine codes. The nine final categories presented in Table 1 were derived inductively from recurring themes that emerged during the review of eligible studies. Once initial themes were identified, all authors, who actively conduct informatics research on clinical informatics, reviewed the categories and reached consensus on their definitions and boundaries. These nine categories— *Outcomes and Effectiveness Evaluations, Secure Messaging Adoption Analyses, User Experience and Engagement Assessments, Message Content Analyses, System Evaluation and Computational Method Development Studies, Generative-AI–Drafted Replies to Patient Messages, Proxy Use and Shared Access Investigations, Health and Technology Literacy Barrier Assessments,* and *Billing and Reimbursement Policy Investigations*—formed the framework for categorizing research themes and synthesizing findings.

**Table 1.** Coding Framework for SM research themes.

| Code Category | Example Codes |
| --- | --- |
| Outcomes and Effectiveness Evaluations | use of SM as an intervention; SM as a predictive tool; evaluation of clinical or behavioral outcomes |
| Secure Messaging Adoption Analyses | SM adoption rates; proportion of SM users among EHR enrollees; usage behavior analysis |
| User Experience and Engagement Assessments | user perspective/satisfaction/preferences; comparative analysis of SM vs. other methods; strategies to improve engagement |
| System Evaluation and Computational Method Development Studies | development of SM-related computational tools; evaluation of SM system functionality; user interface design |
| Generative-AI–Drafted Replies to Patient Messages | evaluation and user perceptions of AI vs clinician response quality; assessment of utilization; prompt engineering and context grounding; disclosure of AI involvement and authorship recognition; identification of risks and safety concerns |
| Message Content Analyses | sentiment analysis; topic or theme analysis; linguistic analysis |
| Proxy Use and Shared Access Investigations | examination of proxy message authorship; issues related to shared access to SM |
| Health and Technology Literacy Barrier Assessments | analysis of patient health literacy and education; technological literacy and associated barriers |
| Billing and Reimbursement Policy Investigations | evaluation of e-visit billing codes; reimbursement-related SM usage trends; financial incentives and barriers |

Two of the three researchers (YG, DH, and YZ) independently coded each article, and discrepancies were resolved through consensus or input from the third reviewer (KZ). A structured coding scheme was developed with open-text fields to capture additional details. We coded research themes using a multi-select field comprising nine inductively derived categories, with the study’s objective recorded verbatim and an “other” option for edge cases. In practice, although this manuscript reports primarily on thematic findings due to the large scale and heterogeneity of included studies, the extraction instrument also captured study design, data scope, data source, and site scope (single-site vs. multi-site), as well as population, clinical domain, and healthcare setting. Outcomes, key results, and author-stated recommendations were recorded in structured free-text fields, with the full instrument available in Supplementary Table S2.

## Data Availability

The data analyzed in this study were derived from previously published literature. Synthesized data supporting the findings of this review are available from the corresponding author upon request.

## ACKNOWLEDGEMENT

The authors received no specific funding or external support for this study.

## CONTRIBUTORSHIP STATEMENT

Y.G. conceived the study, led the literature review, performed screening, data collection, and analysis, and drafted the manuscript. D.H. and Y.Z. contributed to screening and data extraction.

Z.Y. and T.L. contributed to screening. T.L.R. and K.Z. provided methodological guidance and critical revisions. All authors reviewed and approved the final manuscript.

## DATA AVAILABILITY STATEMENT

The data analyzed in this study were derived exclusively from previously published literature. The extracted and synthesized data supporting the findings of this review are available from the corresponding author upon reassonable request.

